# A cross-sectional analysis of self-reported needs and health service utilization among transgender women in Lima, Peru

**DOI:** 10.1101/2022.02.01.22270281

**Authors:** Elizabeth A. Carosella, Leyla Huerta, Jerome T. Galea, Leonid Lecca, Karen Ramos, Giulianna Hernandez, Molly F. Franke, Jesús Peinado

## Abstract

**Purpose:** Globally, transgender women (TGW) experience wide-ranging barriers to health and care, with disproportionately high risks of infectious and chronic diseases. Despite these vulnerabilities, research on access to care for transgender populations in low- and middle-income countries is extremely limited. Furthermore, existing studies have primarily focused on human immunodeficiency virus infection and acquired immune deficiency syndrome (HIV/AIDS), with less emphasis on TGW’s broader health needs. This study analyzed patterns of morbidity and health service uptake among TGW in Lima, Peru. The purpose was to better understand health outreach and service needs to inform targeting and design of future community-level interventions.

**Methods:** This cross-sectional study surveyed a convenience sample of 301 TGW in metropolitan Lima, Peru. Data was collected between September – October 2020. This paper provides descriptive statistics and results of bivariate and multivariable regression models.

**Results:** Health coverage and access to care were suboptimal. Less education and older age were positively associated with illness and HIV and tuberculosis (TB) testing. Gender identity sub-group (i.e., woman, trans or transgender, transsexual, “*transformista*,” “*travesti*,” and other) was associated with HIV testing and pre-exposure prophylaxis (PrEP) usage. Both awareness of and interest in PrEP were low, as was usage among those who were interested in taking PrEP.

**Conclusion:** Future public health efforts should be tailored to meet the diverse needs of TGW, expand TB testing, bridge the gap between PrEP interest and use, and increase insurance coverage and access to trans-friendly services to promote improved health outcomes.

## Introduction

Globally, transgender women (TGW) —individuals born as or assigned male sex at birth who identify as female or another gender that is not male—experience severe and wide-ranging structural, cultural, and interpersonal barriers to access to and utilization of healthcare.^1^ These barriers increase their risk of numerous poor health outcomes, including human immunodeficiency virus infection and acquired immune deficiency syndrome (HIV/AIDS), substance use disorders, violence and injury, mental illness, and other chronic diseases.^2–4^

Through these challenges exist in many parts of the world, Latin America is one of the world’s least hospitable places for TGW, ^5^ with pervasive transphobia, cultural conservativism, and *machismo*,^6,7^ a concept of masculinity characterized by traditional beliefs about gender roles and men’s superior societal status. Despite some recent regulatory improvements, most Latin American countries deny TGW legal recognition of their gender identity, access to gender-affirming care, and numerous other benefits and protections.^8^ Discrimination is widespread, including at health facilities, where providers may refuse treatment or give inadequate care to TGW.^9^ For TGW who migrate to cities where they have limited support,^10^ sex work is often one of the only livelihood options available,^11,12^ increasing the risks for STIs, including HIV/AIDS, and other infectious diseases.^13^

The health needs and healthcare utilization of TGW remain understudied.^4,14^ Less than 0.01% of PubMed articles focus on transgender populations, with most articles from the US.^15^ Data are scarce, in part, because it has historically been common to combine transgender populations with men-who-have-sex-with-men (MSM).^16–18^ Although pre-exposure prophylaxis (PrEP) is one of the greatest advances in HIV prevention, in Peru current levels of PrEP awareness, interest, and utilization among TGW are not well understood. The ImPrEP demonstration study^19^ identified several barriers, including long wait times at dispensary sites due to a lack of doctors needed to prescribe PrEP, lack of information, concern about adverse reactions, and logistical issues that impede PrEP delivery at patient appointments. There is also a lack of data on tuberculosis (TB) risk, despite Peru having the second highest TB incidence in the western hemisphere and being the only Latin American country on the WHO’s top 30 high multidrug resistant TB (MDR-TB) burden countries.^20^ We are not aware of TB incidence data among Peruvian TGW; however, research elsewhere suggests a significant risk.^21^

Another limitation to existing research with TGW is that it largely considers a single, homogenous group. However, there are diverse gender identities within the broader transgender community that impact social perceptions, lived experiences, and behavior trends.^19^ In Peru, commonly used terminology ascribes different characteristics: “*travesti*,” refers to TGW who “utilize female dress full time;” “*transformistas*” alternate between masculine and feminine appearance;^22^ while the umbrella term “trans” is not consistently applied in the literature or in practice.^4^ Nuanced quantitative research on sub-groups could improve targeting of interventions to those at higher risk of poor health or barriers to care.

To inform future clinical and public health outreach and interventions by the nongovernmental organizations Socios En Salud [Partners In Health] and Feminas de Lima, we explored healthcare needs, including health coverage and utilization of HIV and TB testing and gender affirming care, and associated factors, in TGW in Lima, Peru.

## Methods

### Study setting

The exact size of the TGW population in Peru is unknown; there are no national population-level data on transgender health. Research from the US^23,24^ estimates transgender identity prevalence at 0.3% to 0.5%, which, if applied to Peru, suggests that the transgender population overall could be close to 165,000 people. HIV prevalence among Peruvian TGW is 31.8%, in contrast to 10% among MSM,^25^ and 0.3% prevalence in the general population.^26^ Yet, testing among TGW remains low.^27^

In 2016, Peru’s Ministry of Health (MINSA) established a resolution^28^ for STI and HIV infection prevention and control among TGW. MINSA guidelines recommend HIV and syphilis testing on a quarterly basis and outline hepatitis B vaccination, distribution of condoms and lubricant, and the process for hormone therapy for TGW. Only surgeons with specialized training involving transgender patients may administer hormone therapy. In practice, few public facilities provide these services and most transgender-sensitive and gender-affirming healthcare available is accessed through the private sector^26^ or in conjunction with participation in clinical research, especially HIV clinical trials.^29^

Peru’s food and drug administration, *Dirección General de Medicamentos, Insumos y Drogas* (DIGEMID), approved pre-exposure prophylaxis (PrEP) in 2017 following its participation in the landmark iPrEx trial. However, there is a continued lack of national policies and guidelines, which contributes to limited uptake. Mostly, PrEP is accessible to TGW through participation in research, such as the MINSA-sponsored ImPrEP demonstration project^30^ or buying it over the counter.^31^ The organization PrEP Watch estimates that there are only 2,500 – 3,000 PrEP users in Peru as of October 2021.^32^

### Study design and population

We conducted an exploratory cross-sectional study of socio-demographic characteristics and health service utilization among Peruvian TGW in the capital city of Lima, with data collected between September – October 2020. The study population was drawn using a convenience sampling approach, targeting TGW recruited from 21 of 43 districts in Lima. All participants were members of Feminas de Lima, a non-governmental organization comprised of and led by TGW, which is dedicated to transgender health and advocacy.^33^

### Participants and recruitment

Feminas members (N = 311), all of whom identify as part of the TGW community, in participating districts were contacted by phone and invited to participate.

Respondents were at least 18 years old, identified as TGW, and resided in a district of metropolitan Lima. Participants were excluded if they lacked capacity to consent or complete the survey or reported active SARS-CoV-2 symptoms. Peer enumerators - TGW affiliated with Feminas – made in-person visits to each respondent, confirmed fulfillment of criteria, and administered the survey by reading the questions aloud. Ten people either refused to participate or did not respond to phone calls. A total of 301 TGW responded, 46.5% born in Lima and 53.5% originally from other departments. Each respondent gave informed consent.

### Data collection

The survey instrument was developed in Spanish. It was piloted with four Feminas members to test comprehension, time to completion, and overall ease of use before being administered among the full survey sample. Peer enumerators read questions aloud in Spanish and entered responses into a digital form, which staff later uploaded to a central database, which alerted the enumerator if any question was not answered. Surveys took approximately 40 minutes to complete, and there was no missing data.

### Study Measures

Survey questions were based on a prior questionnaire developed by Socios En Salud researchers and tested with Feminas in 2019. Data included demographics, including department of origin (i.e., birthplace, categorized as Lima or outside Lima); age; current residence (house, apartment, room, hostel, no fixed address); highest level of education, and current employment (yes/no). Those currently working could select multiple options for employment type (working for a business, working independently, and sex work). To capture gender identity, we asked participants “how do you define yourself” with response options (Table 1) defined by Feminas based on their knowledge of the TGW community as well as an option to select “other.” The survey included binary response options to the questions “do you suffer from an illness?”; “do you attend a health facility?” and, if so, public or private; “have you heard of PrEP?” and, if so, “would you be interested in PrEP?” and “have you ever taken PrEP?”. There were multiple-choice questions for “how frequently do you get checkups at your health facility?”, “do you have health insurance?”, “have you ever been tested for HIV?”, and “have you ever been tested for TB?” with responses reported in Table 2.

**Table 1:** Socio-demographic Characteristics of a Sample of Transgender Women in Lima, Peru (N = 301)

| Age | Mean ± Standard Deviation<br>n (%) |
| --- | --- |
| Mean | 32.4 ± 8.44 |
| Range | 18 - 64 |
| < 27 years | 73 (24.3%) |
| 27-35 years | 141 (46.8%) |
| > 35 years | 87 (28.9%) |
| **Education** |  |
| ≤ Primary | 19 (6.3%) |
| Secondary | 172 (57.1%) |
| > Primary | 110 (36.5%) |
| **Hometown** |  |
| Lima | 140 (46.5%) |
| Other departments | 161 (53.5%) |
| **Employment** |  |
| Currently Working | 238 (79.1%) |
| *(Of those who responded yes)* |  |
| Work for a business <sup>a</sup> | 59 (19.6%) |
| Work independently <sup>a</sup> | 50 (16.6%) |
| Sex work <sup>a</sup> | 145 (48.2%) |
| **Gender identity** |  |
| Woman | 46 (15.3%) |
| Trans or transgender | 176 (58.5%) |
| Transsexual | 25 (8.3%) |
| *Transformista* | 27 (9.0%) |
| *Travesti* | 21 (7.0%) |
| Other | 6 (2.0%) |

<sup>a</sup> non-exclusive response options

**Table 2:**
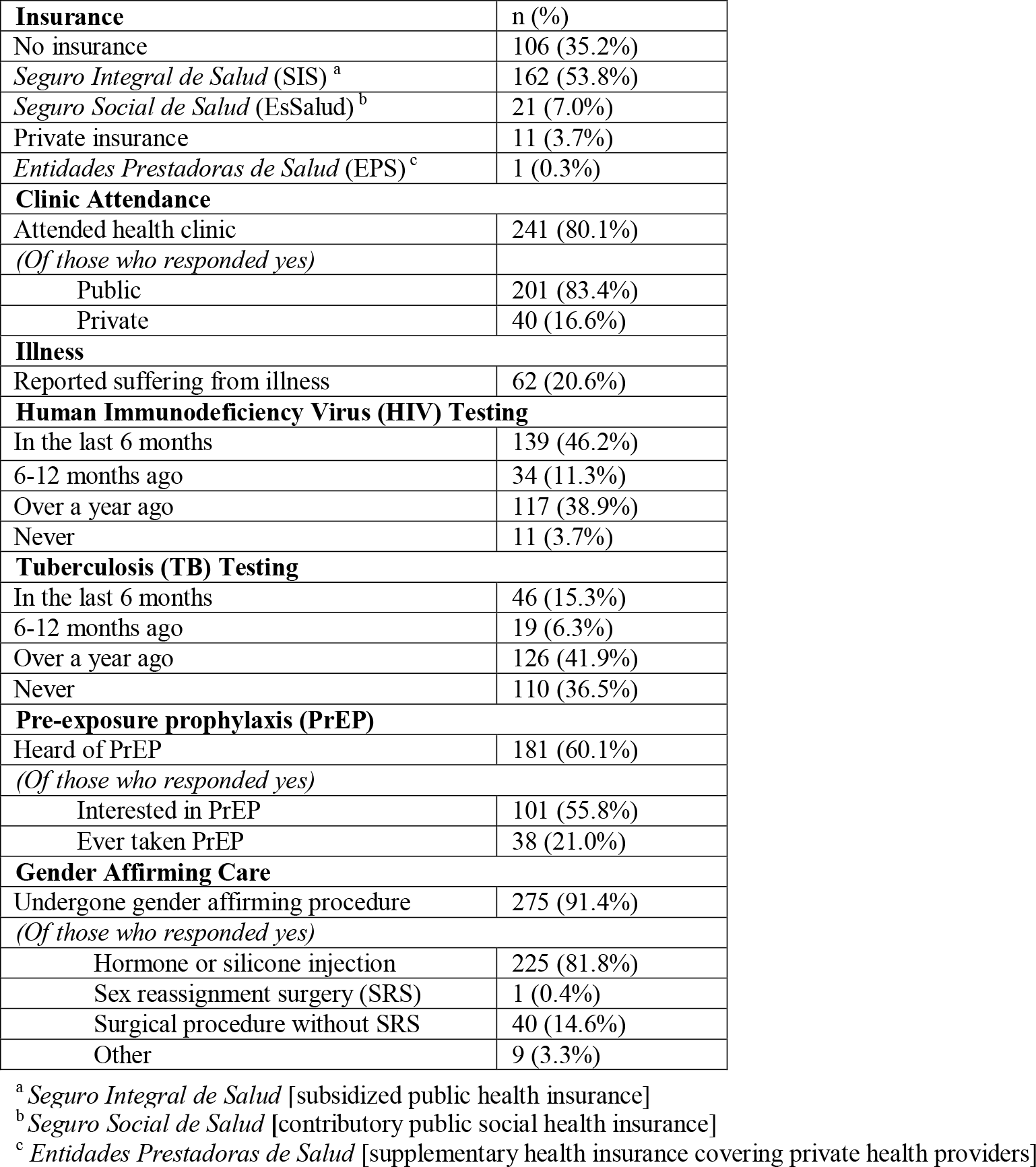
Health and Service Utilization among a Sample of Transgender Women in Lima, Peru (N = 301)

### Ethical considerations

The research protocol and informed consent were approved by the Institutional Review Board of the Peru National Health Institute of Health in June 2020 (OEE-002-20). Participants provided informed consent and we provided them a kit consisting of a face mask, hand sanitizer, condoms, lubricants, and information on where they could access free HIV testing, but no other incentives. The research was completed in accordance with the Declaration of Helsinki as revised in 2013.

### Statistical Analysis

Data were analyzed in Stata 17 (StataCorp LP, College Station, TX, USA). Sample characteristics were described by frequencies and percentages. Age was examined as a categorical variable (i.e., younger than 27 years, 27 – 35, and 36 and older), with upper and lower thresholds corresponding to the highest and lowest quartiles. For gender identity analyses, the self-reported “transgender” category was pooled with “trans,” as Feminas leadership confirmed that these terms are understood synonymously. We evaluated whether these characteristics were associated with binary outcomes for insurance coverage, self-reported illness, regular clinic attendance, TB and HIV testing, and PrEP usage.

Bivariable Poisson regression models with robust error variance were fit to calculate unadjusted prevalence ratios (PRs) and 95% confidence intervals (CIs) for risk factors and six outcome measures: being uninsured, self-reported illness, clinic attendance, TB testing, HIV testing, and PrEP use. The potential risk factors for these outcomes included age group, education level, department of origin, current employment, sex work, and gender identity subgroup. These factors were selected because they were hypothesized to be potential markers of suboptimal health and/or health access within the TGW community in Peru, thereby serving as a means to identify individuals who could benefit from additional support. To calculate adjusted prevalence ratios (aPRs), we then fit a multivariable model for each outcome in which we included all factors of interest (i.e., each factor was adjusted for the others). We largely avoid reference to statistical significance and emphasize CIs to allow analysis of the direction and strength of observed effects.

## Results

### Descriptive Statistics

#### Socio-demographics

Socio-demographic characteristics among 301 respondents are shown in Table 1. Mean age was 32.4 years (range 18-64). 36.5% had completed higher than secondary education. The majority were currently working (79.1%). Sex work was the most common livelihood (48.2%). More than half were born outside Lima (53.5%). Trans or transgender (58.5%) was the most common gender identity, followed by women (15.3%), *transformistas* (9.0%), transsexuals (8.3%), and *travestis* (7.0%).

#### Healthcare utilization

##### Insurance

As described in Table 2, 35.2% of respondents were uninsured. Subsidized public health insurance *Seguro Integral de Salud* (SIS) was the most common (53.8%) insurance option, followed by the contributory public social health insurance (EsSalud) (7%), private insurance (3.7%), and supplementary private insurance (EPS) (0.3%).

##### Clinic Attendance

A high proportion of TGW regularly attended a health clinic (80.1%), with most using public facilities (83.4%).

##### HIV and TB

Most participants reported prior testing for HIV (96.3%), though testing frequency within 12-months was lower (57.5%). A considerable percentage (36.5%) had never been evaluated for TB. Among the total sample, 60.1% were aware of PrEP, of whom 55.8% expressed interest in initiating it (33.6% of the sample); 21.0% (12.6% of the sample) reported they had ever used PrEP.

##### Gender-affirming Care

Respondents commonly reported access to gender affirming procedures (91.4%); among these, most reported non-surgical procedures.

#### Factors associated with health care usage

##### Insurance

As shown in Table 3, older TGW aged 27 – 35 were 38% less likely than TGW under age 27 to be uninsured (aPR for 27 – 35 years = 0.62, 95% CI: 0.43, 0.90). The trend was also observed for TGW over age 35, (aPR for > 35 years = 0.67, 95% CI: 0.43, 1.04) though the CI included 1. Compared with respondents who had higher than secondary education, TGW with primary education or less were 77% more likely to be uninsured (aPR = 1.77, 95% CI: 1.04, 3.03).

**Table 3:** Healthcare Utilization and Access among a Sample of Transgender Women in Lima, Peru - Prevalence Ratios and 95% Confidence Intervals.

|  |  | Uninsured |  | Illness |  | Clinic Attendance |  | Tuberculosis (TB) Testing |  | Human Immunodeficiency Virus (HIV) Testing |  | Pre-exposure prophylaxis (PrEP) Use |  |
| --- | --- | --- | --- | --- | --- | --- | --- | --- | --- | --- | --- | --- | --- |
|  | N = 301 | Unadjusted | Adjusted <sup>a</sup> | Unadjusted | Adjusted <sup>a</sup> | Unadjusted | Adjusted <sup>a</sup> | Unadjusted | Adjusted <sup>a</sup> | Unadjusted | Adjusted <sup>a</sup> | Unadjusted | Adjusted <sup>a</sup> |
| **Age Group** (Reference: < 27 years) |  |  |  |  |  |  |  |  |  |  |  |  |  |
| 27-35 | n = 141<br>(46.8%) | 0.62<br>(0.43, 0.89) | 0.62<br>(0.43, 0.90) | 1.29<br>(0.63, 2.64) | 1.13<br>(0.55, 2.32) | 1.10<br>(0.93, 1.29) | 1.08<br>(0.91, 1.27) | 0.69<br>(0.39, 1.24) | 0.57<br>(0.31, 1.05) | 1.02<br>(0.77, 1.36) | 0.93<br>(0.71, 1.23) | 0.58<br>(0.30, 1.13) | 0.70<br>(0.35, 1.37) |
| > 35 | n = 87<br>(28.9%) | 0.63<br>(0.42, 0.96) | 0.67<br>(0.43, 1.04) | 2.32<br>(1.16, 4.66) | 1.58<br>(0.78, 3.22) | 1.19<br>(1.00, 1.40) | 1.15<br>(0.97, 1.36) | 0.27<br>(0.11, 0.71) | 0.27<br>(0.10, 0.72) | 0.62<br>(0.42, 0.93) | 0.68<br>(0.45, 1.01) | 0.79<br>(0.37, 1.66) | 1.05<br>(0.47, 2.34) |
| **Education** (Reference: > Secondary) |  |  |  |  |  |  |  |  |  |  |  |  |  |
| ≤ Primary | n = 19<br>(6.3%) | 1.75<br>(1.06, 2.90) | 1.77<br>(1.04, 3.03) | 2.59<br>(1.13, 5.93) | 2.61<br>(1.07, 6.40) | 1.05<br>(0.84, 1.32) | 1.05<br>(0.84, 1.32) | 0.26<br>(0.04, 1.78) | 0.29<br>(0.05, 1.89) | 0.65<br>(0.36, 1.20) | 0.71<br>(0.40, 1.24) | 0.55<br>(0.08, 3.69) | 0.44<br>(0.08, 2.54) |
| Secondary | n = 172<br>(57.1%) | 1.09<br>(0.76, 1.56) | 1.07<br>(0.73, 1.56) | 1.79<br>(1.00, 3.21) | 1.50<br>(0.81, 2.78) | 1.03<br>(0.91, 1.16) | 1.00<br>(0.88, 1.14) | 0.56<br>(0.32, 0.98) | 0.61<br>(0.35, 1.06) | 0.66<br>(0.51, 0.85) | 0.70<br>(0.54, 0.90) | 1.16<br>(0.64, 2.10) | 1.20<br>(0.61, 2.38) |
| **Department of Origin** (Reference: Departments other than Lima) |  |  |  |  |  |  |  |  |  |  |  |  |  |
| Lima | n = 140<br>(46.5%) | 0.86<br>(0.62, 1.19) | 0.92<br>(0.65, 1.29) | 1.34<br>(0.83, 2.15) | 1.13<br>(0.70, 1.84) | 1.00<br>(0.89, 1.13) | 0.98<br>(0.87, 1.10) | 0.67<br>(0.37, 1.20) | 0.64<br>(0.36, 1.15) | 0.79<br>(0.61, 1.03) | 0.80<br>(0.62, 1.04) | 0.99<br>(0.55, 1.78) | 0.68<br>(0.36, 1.28) |
| **Employment** (Reference: Unemployed) |  |  |  |  |  |  |  |  |  |  |  |  |  |
| Employed | n = 238<br>(79.1%) | 1.22<br>(0.78, 1.91) | 1.14<br>(0.69, 1.87) | 0.37<br>(0.23, 0.58) | 0.48<br>(0.27, 0.85) | 0.86<br>(0.77, 0.96) | 0.88<br>(0.76, 1.02) | 1.04<br>(0.51, 2.12) | 1.01<br>(0.47, 2.18) | 1.06<br>(0.76, 1.47) | 0.95<br>(0.66, 1.37) | 0.68<br>(0.36, 1.28) | 0.52<br>(0.21, 1.27) |
| **Sex Work** (Reference: Not engaged in sex work) |  |  |  |  |  |  |  |  |  |  |  |  |  |
| Sex work | n = 145<br>(48.2%) | 1.13<br>(0.81, 1.56) | 0.90<br>(0.61, 1.33) | 0.48<br>(0.29, 0.81) | 0.75<br>(0.39, 1.43) | 0.92<br>(0.82, 1.04) | 1.00<br>(0.87, 1.15) | 0.99<br>(0.57, 1.73) | 0.74<br>(0.38, 1.44) | 1.06<br>(0.82, 1.36) | 0.98<br>(0.73, 1.31) | 1.00<br>(0.56, 1.79) | 1.31<br>(0.55, 3.09) |
| **Gender Identity** (Reference: Trans or transgender) |  |  |  |  |  |  |  |  |  |  |  |  |  |
| Woman | n = 46<br>(15.3%) | 1.06<br>(0.70, 1.62) | 1.09<br>(0.72, 1.66) | 0.69<br>(0.31, 1.55) | 0.77<br>(0.35, 1.69) | 0.93<br>(0.77, 1.12) | 0.93<br>(0.77, 1.12) | 1.64<br>(0.85, 3.17) | 1.69<br>(0.89, 3.22) | 1.55<br>(1.19, 2.02) | 1.49<br>(1.16, 1.91) | 1.94<br>(1.00, 3.76) | 2.36<br>(1.15, 4.80) |
| Trans-sexual | n = 25<br>(8.3%) | 0.72<br>(0.35, 1.47) | 0.75<br>(0.36, 1.57) | 1.11<br>(0.48, 2.55) | 0.94<br>(0.44, 2.02) | 1.07<br>(0.90, 1.28) | 1.05<br>(0.88, 1.24) | 1.57<br>(0.66, 3.71) | 1.66<br>(0.72, 3.80) | 1.09<br>(0.69, 1.72) | 1.09<br>(0.69, 1.73) | 1.85<br>(0.74, 4.64) | 1.62<br>(0.66, 4.02) |
| *Transformista* | n = 27<br>(9.0%) | 0.59<br>(0.21, 1.64) | 0.62<br>(0.23, 1.67) | 1.45<br>(0.60, 3.52) | 1.01<br>(0.39, 2.59) | 1.15<br>(0.98, 1.35) | 1.08<br>(0.91, 1.27) | 1.03<br>(0.27, 3.92) | 1.09<br>(0.33, 3.60) | 0.98<br>(0.52, 1.83) | 0.98<br>(0.49, 1.94) | undefined | undefined |
| *Travesti* | n = 21<br>(7.0%) | 0.81<br>(0.38, 1.73) | 0.76<br>(0.38, 1.53) | 1.80<br>(0.88, 3.66) | 1.21<br>(0.60, 2.44) | 1.02<br>(0.81, 1.28) | 0.99<br>(0.78, 1.22) | undefined | undefined | 0.40<br>(0.14, 1.14) | 0.50<br>(0.17, 1.44) | 0.67<br>(0.10, 4.46) | 0.51<br>(0.09, 2.90) |
| Other | n = 6<br>(2.0%) | 1.37<br>(0.50, 3.74) | 1.15<br>(0.45, 2.90) | undefined | undefined | 0.62<br>(0.23, 1.65) | 0.62<br>(0.24, 1.61) | 1.80<br>(0.32, 10.27) | 1.49<br>(0.35, 6.37) | 1.14<br>(0.42, 3.08) | 1.00<br>(0.38, 2.70) | 6.00<br>(3.93, 9.16) | 5.64<br>(2.75, 11.58) |

<sup>a</sup> Covariates included age group, education level (primary or less, secondary, or higher than secondary), department of origin (Lima or other department of origin), current employment (yes/no), sex work (yes/no), and gender identity subgroup

##### Illness

TGW with primary education or less had higher prevalence of self-reported illness (Apr = 2.61, 95% CI: 1.07, 6.40). Employed TGW had a lower prevalence of self-reported illness (aPR = 0.48, 95% CI: 0.27, 0.85). Older TGW were more likely to report illness though CIs for these estimates were wider and included one (Table 3).

##### Clinic Attendance

Older TGW were more likely to report clinic attendance (aPR for 27-35 years = 1.08, 95% CI: 0.91, 1.27; aPR for > 35 years = 1.15, 95% CI: 0.97, 1.36). Employed TGW had lower clinic attendance (aPR = 0.88, 95% CI: 0.76, 1.02). In an a posteriori analysis, we examined whether sex work could moderate the relationship between employment and illness or clinic attendance but did not find statistical evidence of this (p = 0.31 for interaction).

##### TB Testing

Older TGW were less likely to have ever had a TB test (aPR for 27 – 35 years = 0.57, 95% CI: 0.31, 1.05; aPR for > 35 years = 0.27, 95% CI: 0.10, 0.72). This was also true of those with lower levels of education (aPR for primary education or less = 0.29, 95% CI: 0.05, 1.89; aPR for secondary education = 0.61, 95% CI: 0.35, 1.06) though CIs included 1. Employment did not appear to impact TB testing (aPR = 1.01, 95% CI: 0.47, 2.18).

##### HIV Testing

Older TGW, particularly those over age 35, were less likely to have ever had an HIV test (aPR for those > 35 years = 0.68, 95% CI: 0.45, 1.01). Older age also was inversely associated with sex work (aPR for 27 – 35 years = 0.69, 95% CI: 0.55, 0.87; aPR for those > 35 years = 0.44, 95% CI: 0.31, 0.64). As with TB, lower levels of education were associated with lower likelihood of HIV testing (aPR for primary education or less = 0.71, 95% CI: 0.40, 1.24; aPR for secondary education = 0.70, 95% CI: 0.54, 0.90). Again, employment did not appear to be associated with HIV testing (aPR = 0.95, 95% CI: 0.66, 1.37). TGW who identified as women were more likely to test for HIV (aPR = 1.49, 95% CI: 1.16, 1.91).

##### PrEP and gender affirming care

Compared to those who identified as trans or transgender TGW who identified as women were more likely to use PrEP (aPR = 2.36, 95% CI: 1.15, 4.80) and report having had surgical procedures: PR: 1.73 (0.92, 3.24). Those who indicated their gender identity as “other” were also more likely to use PrEP (aPR = 5.64, 95% CI: 2.75, 11.58). We did not find strong evidence that any other factors were associated with PrEP use or gender affirming care.

## Discussion

This study identified a diverse population with varying health needs and service utilization patterns. Overall, TGW had suboptimal levels of health coverage and care, with less education and older age as important risk factors for many outcomes. In the first study to quantitatively examine health utilization by gender identity subgroups in Peru, we also found that TGW who identified as women had higher HIV testing and PrEP uptake compared to those who identified as trans or transgender.

Nearly one in five respondents reported that they did not receive facility-based healthcare. This may be due to a combination of factors that contribute to poor linkages to and retention in healthcare.^34^ Over one-third of our sample did not have insurance of any kind, with the lowest levels of insurance among younger and less educated TGW. In Peru, national ID cards typically do not reflect TGW’s true gender identity,^35^ which can obstruct administrative processes. Many TGW either lack ID cards entirely or do not use them for fear of being subjected to ridicule or abuse, including in health centers.^35^ Because national ID cards are required for Peruvians to enroll in the SIS, despite regulatory changes in recent years to loosen restrictions for changing gender on national IDs, this remains a barrier to healthcare access.

The lack of differentiated services or trans-friendly spaces within Peru’s health system is likely another important barrier.^26^ Gender-affirming healthcare is rarely provided in public facilities and we found a high prevalence of non-surgical gender affirming care practices-mainly, hormone injections and silicone implants, which are often obtained outside the health system.^10,11,22^ Previous studies found that TGW may self-administer treatments or rely on medically-unqualified acquaintances.^11,22^ Medical providers and staff may lack training and cultural competency to provide quality services to TGW.^36,37^ The expectation of hostile, discriminatory, or substandard treatment may deter even those who have public insurance from seeking facility-based care. To date most scholarship has focused on patient perspectives, and we call for more research on providers’ knowledge, attitudes, and other supply-side barriers to care facing TGW.

Though HIV testing was generally high, older and less educated TGW were less likely to be tested. Older TGW may have had less exposure or access to preventive testing, free anti-retroviral treatment, and informational and anti-stigma campaigns, which have become more common over time. Moreover, older age was negatively associated with sex work. We hypothesize that less frequent sex work among older women may reduce older TGW’s HIV risk or risk perception, leading them to test less frequently.

In contrast with HIV testing, TB testing was low with over one in three TGW never tested. Given Peru’s high national TB burden, TGW may face a disproportionately high risk of infection due to HIV and a tendency to cluster in communal homes. Stigma and discrimination may limit access to housing: nearly half of participants lived in shared housing or hostels in the few districts where they can secure housing.^33,38^

Given that this study’s overarching goal was to help inform and appropriately target future health outreach and interventions, we assessed patterns among different gender identity subgroups to pinpoint those that may be most in need of services. This analysis builds on prior qualitative research about the varied experiences of different gender identity subgroups within Peru’s TGW community and existing quantitative research with TGW that largely assesses them as a homogenous group. We found that TGW who identify as women were 49% more likely to have been tested for HIV compared to those who identify as transgender.

A sub-analysis found that women had nearly twice the prevalence of surgical procedures compared to those who identified as trans or transgender. Together with findings on testing rates, this might reflect that having a physical appearance that aligns with a traditional binary gender reduces friction in TGWs’ interaction with the health system and facilitates access. More qualitative research on how different gender identities impact the testing and broader healthcare utilization could better illuminate this finding. Note that this research was exclusive to TGW, and there is little research available on Peruvian transgender men or non-binary populations.

About two in five TGW (39.9%) had never heard of PrEP, and there was a large gap between those who expressed interest in PrEP (55.8%) and those who reported ever using it (21.0%). Overall, prevalence of PrEP usage was just 12.6%. Prior research highlights several barriers: limited perceived benefits, clinicians’ disregard of side effects, anticipated out-of-pocket costs, and concerns that PrEP would disincentivize condom usage among PrEP users.^29,39^ Mistrust of medical institutions, doubts about safety and efficacy, suspicion of actors offering free PrEP, and feelings of exploitation in the face of persistent barriers to PrEP access outside of research environments are also common.^29^ Further research among those who report interest but not uptake could disentangle supply-versus demand-side factors and inform strategies to increase PrEP knowledge and demand. Looking forward, the US FDA approved long-acting injectable PrEP-cabotegravir (CAB - LA)^40,41^ in December 2021,^42^ which may reduce barriers to dosing, adherence, and stigma related to daily oral medication.

At the time of study, authors were unaware of any community-level interventions that address broader health issues among Lima’s TGW. Existing services tailored for TGW have been secured for national HIV surveillance and HIV prevention clinical trials, as well as Ministry of Health screenings, which require going to a health facility. Along with existing HIV programs, there may be opportunities to build on COVID-19 testing strategies and infrastructure that focus on extending health services that reach TGW where they are.

## Limitations

This was an exploratory analysis aiming to identify subgroups of TGW who could be prioritized for interventions to improve access to care. We did not adjust for multiple comparisons because the goal of our analysis was not to identify singular statistically significant associations but rather to identify meaningful patterns of healthcare utilization. There is potential for selection bias; however, we expect that this is limited given Feminas’ extensive network and the high participation rate. The study relied on self-reporting, which could introduce bias, the direction of which may be difficult to predict. Furthermore, while a strength of this study was the participation of 301 TGW respondents, small sample sizes may influence some results such as those for gender identity sub-groups where, for example, just six participants reported “other” gender identities (see Table 3).

Finally, meanings associated with gender identity labels are evolving; certain terms, like “transsexual,” become less commonly used and others like “trans” begin to subsume previous catch-all terms like “*travesti*.” This may limit interpretation or generalizability of associations with specific gender identity. The use of the Feminas peer promoter network to identify study participants and implement the survey assured that respondents’ gender identities fell within the umbrella of TGW. However, we recognize that identity is often fluid and part of a transition process, and that respondents may have been at different stages. Future surveys may be more inclusive by incorporating additional options and utilizing a “check all that apply” approach to gender identity collection.

## Conclusion

We identified gaps in health coverage and services among Peruvian TGW. Outreach and interventions should aim to expand insurance coverage and access to trans-friendly services for TGW who do not seek facility-based care, particularly younger TGW. While HIV testing rates were high, PrEP uptake remains suboptimal. Further efforts are needed to bridge the gap between interest and usage.

Finally, TB research and services should be prioritized, given the paucity of data and the potential for this population to be disproportionately affected.

## Data Availability

All data produced in the present study are available upon reasonable request to the authors

## Acknowledgements

We are grateful to the study participants for sharing their experiences. Socios En Salud Peru developed the survey, collected the data, and analyzed findings. Feminas co-designed the survey, supplied peer community health workers, and contributed to the analysis.

## Authorship Contribution Statement

**Elizabeth A. Carosella:** Methodology, data curation, formal analysis, writing – original draft. **Leyla Huerta:** Conceptualization, investigation, resources, validation. **Jerome T. Galea**: writing – review & editing. **Leonid Lecca**: Conceptualization, resources. **Karen Ramos**: Investigation, supervision. **Giulianna Hernandez**: Investigation, supervision. **Molly F. Franke**: Conceptualization, methodology, validation, writing – review & editing. **Jesus Peinado**: Conceptualization, methodology, validation, supervision, writing – review & editing.

